# State heterogeneity of human mobility and COVID-19 epidemics in the European Union

**DOI:** 10.1101/2020.06.10.20127530

**Authors:** Xiaoling Yuan, Kun Hu, Jie Xu, Xuchen Zhang, Wei Bao, Charles F. Lynch, Lanjing Zhang

## Abstract

Human mobility was associated with epidemic changes of coronavirus disease 2019 (COVID-19) in China, where strict public health interventions reduced human mobility and COVID-19 epidemics. But its association with COVID-19 epidemics in the European Union (EU) is unclear. In this quasi-experimental study, we modelled the temporal trends in human mobility and epidemics of COVID-19 in the 27 EU states between January 15 and May 9, 2020. COVID-19 and human mobility had 3 trend-segments, including an upward trend in COVID-19 daily incidence and a downward trend in most human mobilities in the middle segment. Compared with the EU states farther from Italy, the state-wide lockdown dates were more likely linked to turning points of human mobilities in the EU states closer to Italy, which were also more likely linked to second turning points of COVID-19 epidemics. Among the examined human mobilities, the second turning points in driving mobility and the first turning points in parks mobility were the best factors that connected lockdown dates and COVID-19 epidemics in the EU states closer to Italy. Our findings highlight the state- and mobility-heterogeneity in the associations of public health interventions and human mobility with changes of COVID-19 epidemics in the EU states.

## Introduction

As of May 9, 2020, the coronavirus disease 2019 (COVID-19) affected more than 3,855,809 people in the world.^1^ Strict public health interventions, including cordon sanitaire, have reduced human mobility in China,^2^ which was shown to link to a reduction of COVID-19 epidemics. However, the changes of human mobility in the European Union (EU) states during COVID-19 pandemic have been largely unknown as their associated factors and links to COVID-19 have not yet been explored. We therefore conducted a quasi-experimental study to assess these associated factors and their associations with human mobility.

## Methods

In this quasi-experimental study, we extracted the data of laboratory-confirmed COVID-19 cases from the European Centre for Disease Prevention and Control,^3^ which were reported by the respective member states of the EU during January 1, 2020 to May 9, 2020. The daily incidence was calculated using the denominator of states’ populations, which were sourced from Eurostat. The 26 non-Italy EU states were classified into 3 groups based their neighboring relationship to Italy, including group 1 (Austria [AT], Croatia [HR], France [FR] and Slovenia [SI]), group 2 (Belgium [BE], Bulgaria [BG], Czechia [CZ], Germany [DE], Greece [GR], Luxembourg [LU], Netherlands [NL] and Spain [ES]) and group 3 (the rest).

We extracted the Google mobility and Apple mobility data from their respective websites.^4,5^ Both Google mobility and Apple mobility data were based on aggregated global position system (GPS) data, and represented the percentage change relative to the human mobility at a preset baseline date. Specifically, the Google mobility data were derived from the human mobility at locations including parks, grocery store and pharmacy, retail and recreation business, workplace and transit station.^4^ It has been used in predicting COVID-19 trends.^6^ The Apple mobility data were derived from individuals’ mobility of driving, walking and transit.^5^ Therefore, the Google mobility data are more focused on the aggregated human mobility in a given location, while the Apple mobility data are more focused on individual mobility activities. They represented different approaches to human mobility. All data were de-identified and publicly available. A review by the institutional review board is thus exempt (category 4).

The dates of implementing state-wide lockdown and social distancing, and lifting lockdown-bans in a state were extracted from the Wikipedia page,^7^ and Institute for Health Metrics and Evaluation, if needed.^8^ The simulation study was conducted based on the coefficients computed using log-linear modelling of daily incidence. Specifically, the percentage changes equaled the exponential of the days of moving earlier (-d) multiplied by the coefficient (β) (i.e. change = e^(-d x β)).

Statistical analyses were conducted using Stata (version 15). We used piecewise log-linear or ordinal least square regression models to identify the turning points of the daily new cases or human mobility. Two turning points (i.e. 3 segments) were assumed in each modelling. The locally weighted scatter smoothing (LOWESS) algorithm was used to smooth daily incidence and mobility trajectories.^9,10^ It is noteworthy that the daily incidence and its changes are proportional to those of daily new cases since the population of a given state remained relatively constant during the study period. Pearson’s correlation and ordinal least square analyses were conducted to assess the potential link to turning points. All *p* values were 2-sided. A *p*<0.05 was considered statistically significant.

## Results

There were 982,332 laboratory-confirmed COVID-19 cases in the 27 EU states (median 7,896, interquartile 1,689 to 25,702 for individual states) from Jan. 1 to May 9, 2020 (**Figure 1 and Supplementary Figure 1**). Nearly all of the EU states had 3 segments of trends in COVID-19 incidence (**Supplementary Table 1**). The piecewise log-linear models show that the COVID-19 daily growth rate in the first segment of Italy was 7.8%, and statistically indifferent from those in the second segments of all other state groups (**STable 2**), suggesting a delayed growth in non-Italy EU states. Interestingly, only the states in group 1 had a much faster decreasing rate than Italy (**Supplementary Table 2**). Among the 27 states, 21 implemented a state-wide lockdown policy, and 25 implemented social distance restrictions. The daily new cases overall had a median increase rate of 8.8% per day (interquartile 5.7-10.5%), which peaked at 67 per million (interquartile 27-118), and then decreased at a median rate of 1.9% (interquartile 3.0-0.0%). The median duration of the second (increasing) segment was 27 days (interquartile 24-35).

**Figure 1.**
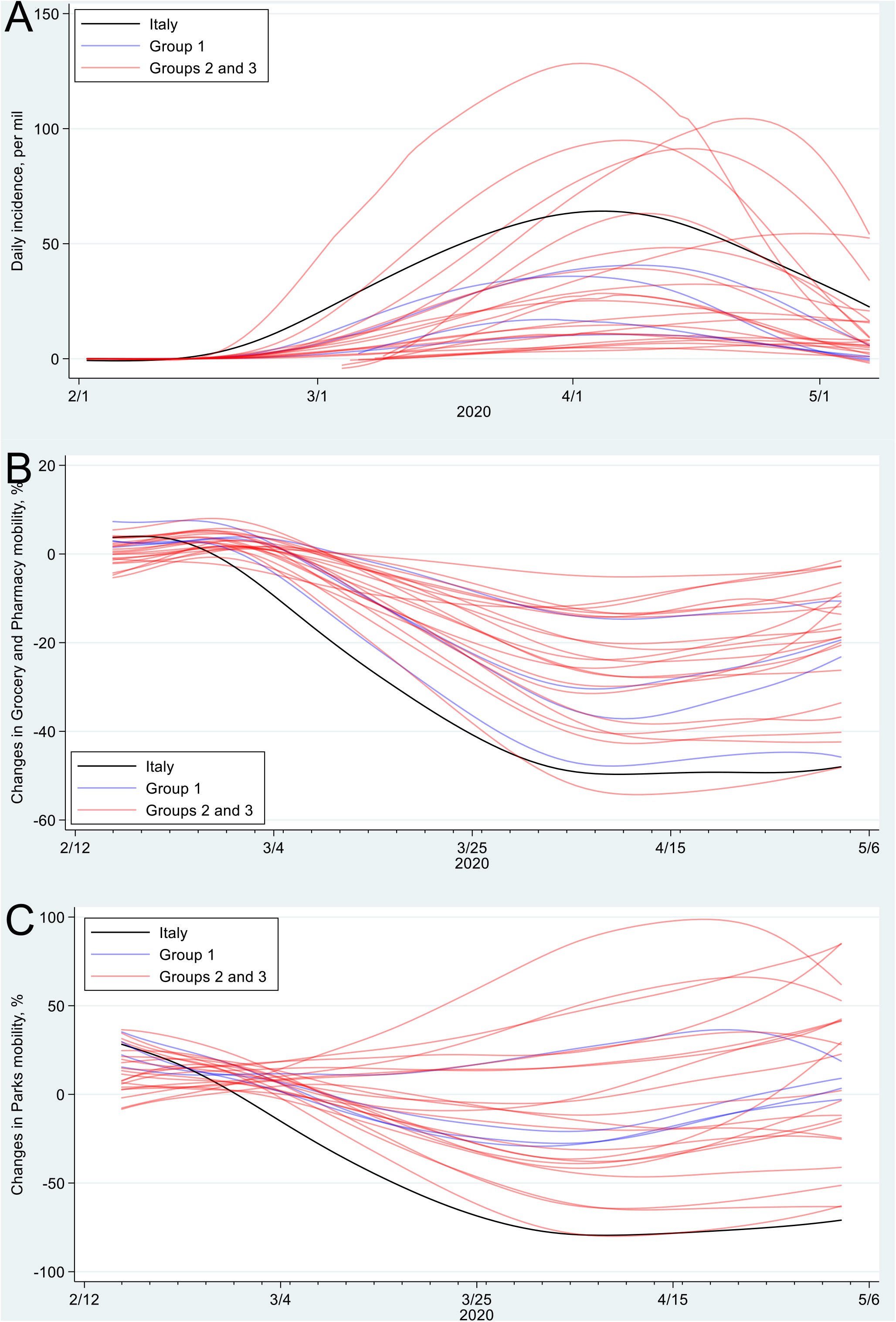
**A**. The daily incidence trends of COVID-19 in the 27 European Union states. **B&C**. The percentage changes of grocery and pharmacy mobility and those of parks mobility in the 27 European Union states. The state groups were classified according to their neighboring relationship to Italy.

Among Google human mobility trends in 26 states (all EU states except Cyprus), residential mobility increased and the mobility of retail and recreation, parks, grocery and pharmacy, workplace, and transit station decreased during their second segments and increased afterward (**Supplementary Table 3**). Among Apple mobility trends in 25 states (all EU states except Cyprus and Malta), the mobilities of walking, driving and transit also decreased during their second segments that were followed by an upward trend (**Supplementary Table 2**).

The associations of second turning points in some mobilities with those in daily new cases and incidence were stronger in the states closer to Italy (**Figures 2 A&B**), with group 1 having the strongest association. The second turning points in most of the tested human mobilities were linked to those in COVID-19 epidemics in the states immediately next to Italy (group 1), while the associations were less strong in the states farther from Italy. The associations of first turning points in parks mobility with those in daily new cases and incidence were stronger in the states closer to Italy (**Figures 2 C&D**). The R-squared (R^2^), as a measure of likelihood of association, were the largest in group 1 and smallest in group 3 in some human mobilities. It suggests the likelihood of such an association was higher in the states that were closer to Italy.

**Figure 2.**
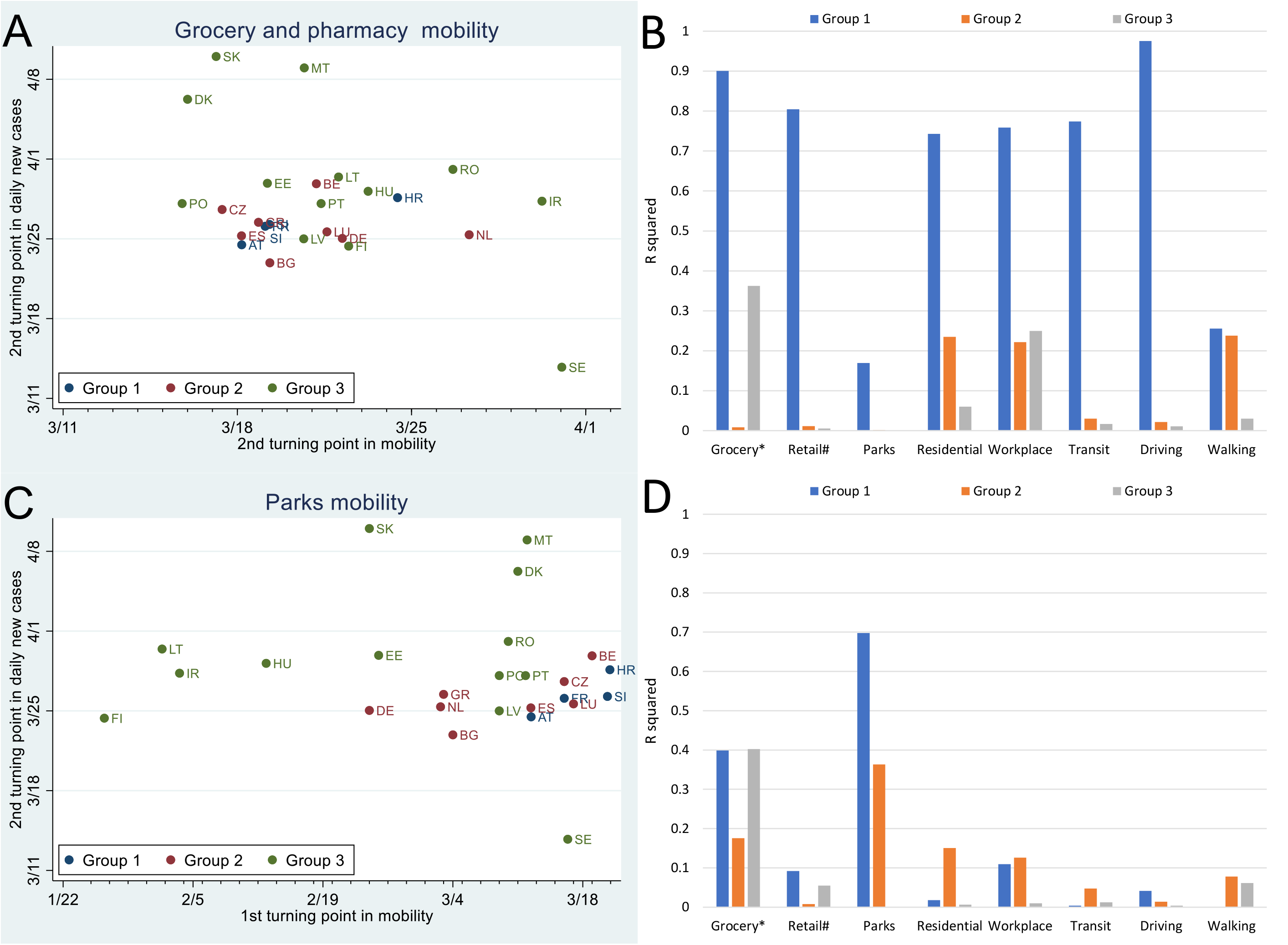
**A.& B**. The associations of second turning points in mobility with second turning points in daily new cases were stronger in the states closer to Italy (the p-value for grocery and pharmacy mobility was 0.012 in group 1). **C**.**&D**. The associations of first turning points in mobility with second turning points in daily new cases showed a similar pattern (the p-value for parks mobility were 0.121 in Group 1 and 0.106 in Group 2). The larger R-squared values indicate stronger associations. The state groups were classified according to their neighboring relationship to Italy. #, also included pharmacy mobility; *, also included recreation mobility.

We then explored the potential factors that might be linked to the turning points of human mobilities. The lockdown dates seemed associated with second turning points in walking mobility (R^2^=0.91, *p*=0.09) in state group 1 (**Figure 3**), while they were strongly associated with second turning points in the mobilities of residential (*p*=0.02), transit (*p*=0.005) and driving (*p*=0.01) in state group 2. Interestingly, the associations of lockdown dates (in 2020) with first turning points in the mobilities of grocery and pharmacy, retail and recreation, and parks appeared to be stronger in the states closer to Italy (*p*=0.004 for grocery and pharmacy mobility in group 1). The dates of implementing social distancing were not associated with the second turning points of human mobilities (**Supplementary Figure 2**). However, their associations with the mobilities of grocery and pharmacy, retail and recreation and transit were stronger in the states closer to Italy.

**Figure 3.**
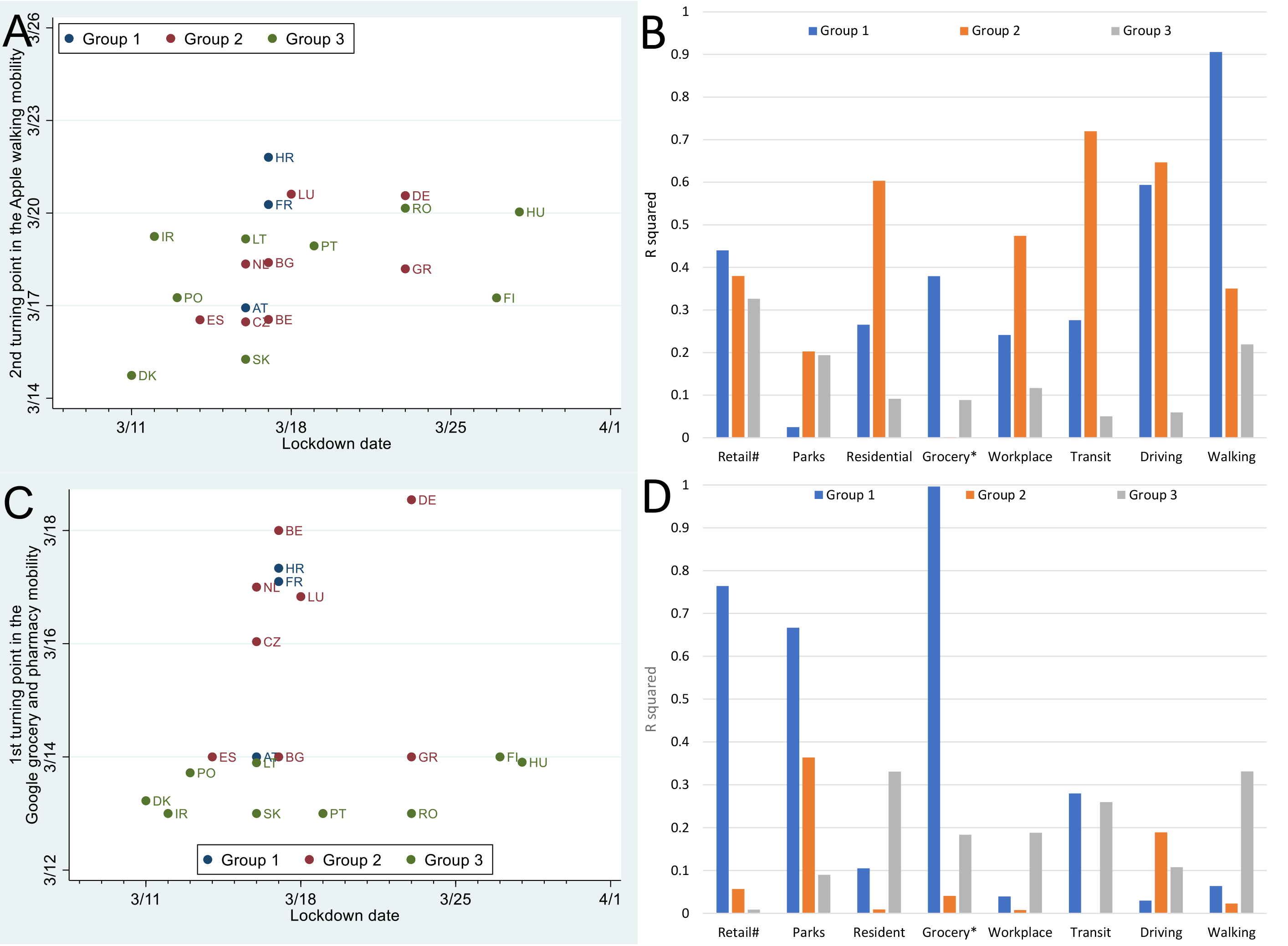
**A.& B**. The associations of lockdown dates (in 2020) with second turning points in the mobilities of retail and recreation, and walking were stronger in the states closer to Italy (the p-value for walking was 0.09 in group 1), while those with second turning points in the mobilities of residential (*p*=0.02), transit (*p*=0.005) and driving (*p*=0.01) were strongest among the states in group 2. **C**.**& D**. The associations of lockdown dates (in 2020) with first turning points in mobility were stronger in the states closer to Italy (the p-value for grocery and pharmacy mobility was 0.004 in group 1). The larger R-squared values indicate stronger associations. The state groups were classified according to their neighboring relationship to Italy. #, also included pharmacy mobility; *, also included recreation mobility.

According to the R^2^ values, the second turning points in driving mobility and the first turning points in parks mobility were the best mediators among the second and first turning points in various mobilities, respectively, for connecting public health intervention (lockdown) date and COVID-19 daily incidence in group 1 states. Specifically, the lockdown dates were linked to the second turning points of driving mobility in group 1 states (R^2^=0.59), which was in turn linked to the daily new cases of COVID-19 (R^2^=0.98). The lockdown dates were also linked to the first turning points of parks mobility in group 1 states (R^2^=0.67), which was then linked to the daily new cases of COVID-19 (R^2^=0.70).

The simulation study shows that the peak daily incidence of COVID-19 would have been greatly decreased had the peak/turning points been reached days earlier (**Figure 4**). For example, if the peak-incidence date had been moved to 7 days earlier, there would have been a 20-80% reduction in peak daily incidence. Such a reduction would have translated to 66 cases/million in Luxembourg (from 374 cases/million).

**Figure 4.**
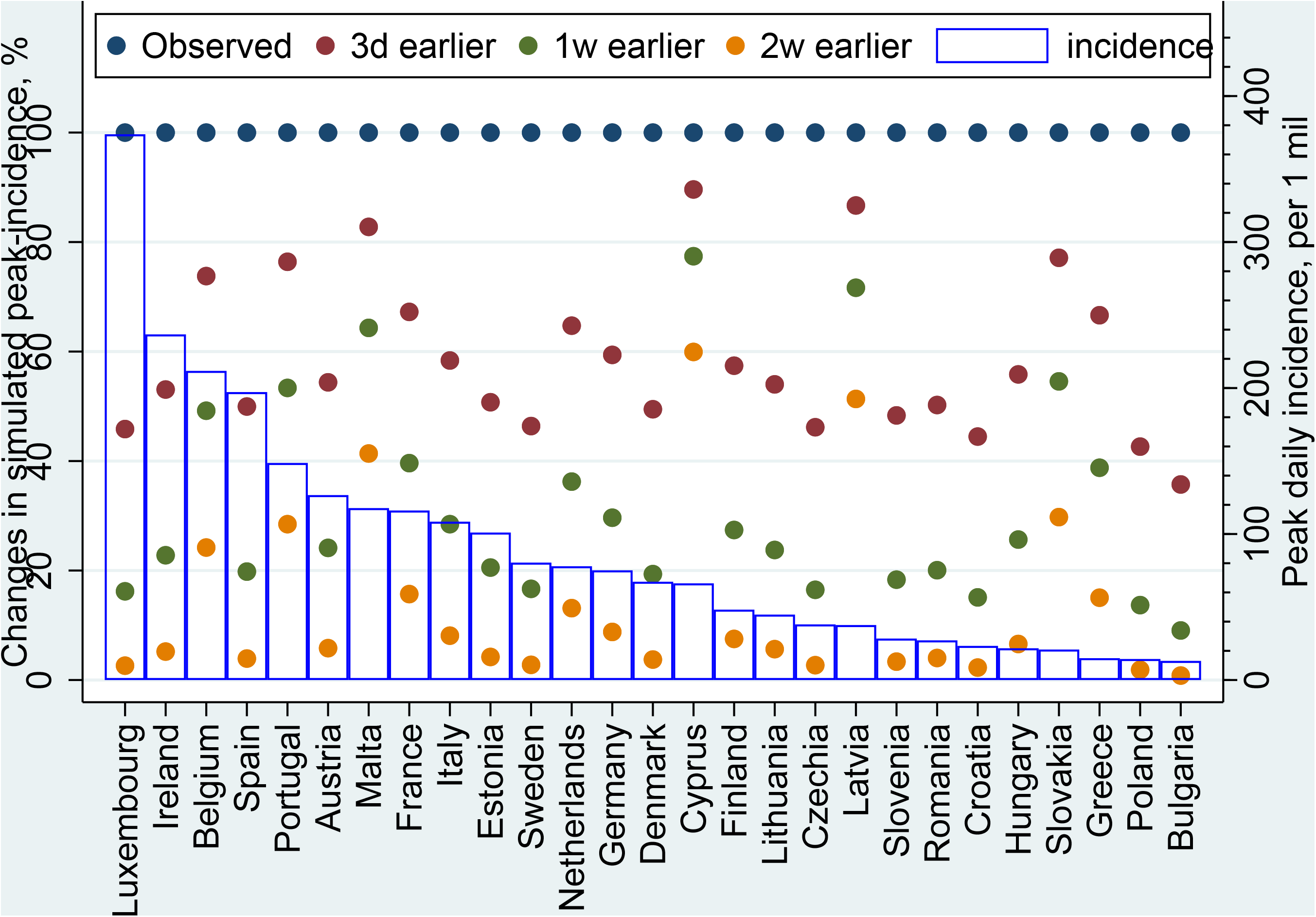
Simulated changes of COVID-19 daily incidence in the European Union states. The scattered dots show the simulated percentage changes of daily incidence by states under the scenarios when the peaks of daily COVID-19 incidence were reached 3 days (3d), 1 week (1w) and 2 weeks (2w) earlier. The daily incidence (empty bar, laboratory-confirmed COVID-19 cases per million) is also shown.

## Discussion

This quasi-experimental study shows state heterogeneity in the association of human mobilities with COVID-19 epidemics in the EU states, and in the lockdown dates with human mobilities. These associations appear to be stronger in the states closer to the COVID-19 epicenter (Italy at the time) than the states farther. There was also modality heterogeneity of human mobility in these associations. The second turning points in driving mobility and the first turning points in parks mobility were the best mediators that connected public health intervention (lockdown) date and COVID-19 daily incidence in group 1 states.

The turning points of some human mobilities were associated with those of COVID-19 epidemics in the EU states in the group 1, but not in other groups. Thus, the neighboring relationship to the COVID-19 epicenter may determine or influence the associations of the lockdown dates with turning points in human mobility in EU states, and subsequently those with COVID-19 epidemics. In other words, there was considerable state heterogeneity in the COVID-19 epidemics of EU states, which seemed linked to human mobility and indirectly to the lockdown dates. Therefore, these associations may not be significant in all states combined due to the “dilution” from the non-group 1 states. Indeed, a previous report using susceptible-infected-recovered and machine learning (gradient-boosted trees) models showed that changes of social distancing explained 46% of the variances in COVID-19 transmission rate.^11^ Despite the difference in study design and time-frame of interest, the state heterogeneity shown by this study may in part explain their findings. Furthermore, the state heterogeneity should be considered when implementing public health interventions. Perhaps the states closer to the epicenter state should have considered implementing public health interventions earlier and stricter than other states in the affected continent. However, when and how the states farther away from the epicenter should implement these interventions are not clear and warrant further evaluation.

The modality difference of human mobility in the COVID-19 were rarely examined in previous studies. Most of the previous studies on human mobility and COVID-19 used migration mobility data.^2,12,13^ One study described the temporal trends of mobile device-based mobility (Safegraph) in 4 U.S. metropolitan cities, but did not model any turning points of mobility trends.^14^ Indeed, GPS-based individual mobility data were provided by Google and Apple only after April 2020.^4,5^ The Google and Apple mobility data were based on observations at locations (e.g. residential and work places) and moving modes (i.e. walking, driving and transit), respectively. Such a difference may partially explain why the best interlink factors between lockdown dates and COVID-19 daily incidence were the second turning points in driving mobility (Apple), but the first turning points in parks mobility (Google). The reason for this may be that state-wide lockdown was associated with the bottom in the trends of individual-based driving mobility (Apple data), and with the first turning points in the trends of location-based parks mobility (Google data), while both turning points were linked to the changes in daily incidence. Therefore, policy makers may consider using the first turning point of parks mobility and the bottom/second turning point of driving mobility to monitor the effects of state-wide lockdowns and predict the peak points of COVID-19 daily incidence. A possible cutoff of the lowest driving mobility would be approximately 25% of the baseline mobility, which was the one observed in this study.

Some of this study’s strengths are noteworthy. First, this study was focused on the state heterogeneity in COVID-19 epidemics and their associated factors in EU where less strict public health interventions were implemented for COVID-19 than those in China. We show that the EU states closer to the epicenter (Italy in the EU) were more likely to observe the links between lockdown and human mobility changes, and subsequently the changes in COVID-19 epidemic. Second, we comprehensively analyzed the associations of the GPS-based mobilities with COVID-19 epidemics in the EU states, including location-based Google and individual activity-based Apple mobility data. The previous reports on COVID-19 have used Google and Apple mobilities,^11^ or human migration mobility in Europe, the USA or China.^2,14,15^ However, to our knowledge they have not examined the association of turning points of these mobilities with the changes in both public health intervention and COVID-19 epidemics. Third, this study provides early evidence on whether and how human mobilities were associated with COVID-19 epidemics under less strict state-wide lockdown. Similarly, the association of human migration mobility with COVID-19 differed by the distance to its epicenter (Wuhan at the time) in China,^2^ where the public health interventions were much stricter than the rest of the world and may not be applicable to other states.

A limitation of this study is that some non-EU states in Europe were not analyzed, including the United Kingdom and Switzerland, while their human mobility might also be associated with COVID-19 epidemics. Moreover, the state heterogeneity among the EU states may not be generalizable to other continents because of the unique cultures and geopolitical systems in the EU. China, for example, implemented very strict public health interventions and may not experience significant providence-heterogeneity in the associations of public health interventions with COVID-19 epidemics. Furthermore, the mobility data of Cyprus were not available, and could not be reliably analyzed. Finally, the sensitivity and specificity of COVID-19 tests used in each state might differ, and lead to some variances in the (daily) incidence of COVID-19. However, all clinical tests on the EU markets must have gone through regulatory reviews by the European Medicine Agency, and should have considerably similar sensitivity and specificity. The single-market policy in the EU also reduced differences in the tests’ availability among all EU states.^16^ The variances in COVID-19 test performance thus were minimal in the EU states.

In summary, we showed the heterogeneity of state and modality of human-mobility in COVID-19 epidemics and their associated factors in the EU states. We also characterized the trends in COVID-19 epidemics and human mobilities across the EU states. These findings may help choose the best timing for public health interventions, likely based on the state’s neighboring relationship with the epicenter during the COVID-19 pandemic. Future works should be focused on the factors linked to the trends of COVID-19 epidemics in the states farther from the epicenter.

## Statement

### Authors’ contributions

KH, XY, JX and LZ designed the study, KH, XY and LZ extracted and analyzed the data, KH and XY wrote the first draft of the manuscript and all authors edited the manuscript. The final manuscript was approved by all authors.

No conflict of interest is declared by any of the authors. No funding sources are reported.

### Data Availability

All data are readily available at respective websites.

## Data Availability

Data Availability: All data are readily available at respective websites.

## References

1 WHO. Coronavirus disease (COVID-2019) situation reports, <https://www.who.int/emergencies/diseases/novel-coronavirus-2019/situation-reports/> (2020).

2 Kraemer, M. U. G. et al.. The effect of human mobility and control measures on the COVID-19 epidemic in China. Science 368, 493–497, doi:10.1126/science.abb4218 (2020).

3 Ritchie, H. Coronavirus Source Data, <https://ourworldindata.org/coronavirus-source-data> (2020).

4 Google. COVID-19 Community Mobility Reports, <https://www.google.com/covid19/mobility/> (2020).

5 Apple. Apple Maps Mobility Trends Reports, <https://www.apple.com/covid19/mobility> (2020).

6 Ayyoubzadeh, S. M., Ayyoubzadeh, S. M., Zahedi, H., Ahmadi, M. & S, R. N. K. Predicting COVID-19 incidence using Google Trends and data mining techniques: A pilot study in Iran. JMIR public health and surveillance, doi:10.2196/18828 (2020).

7 Wikipedia. COVID-19 pandemic lockdowns, <https://en.wikipedia.org/wiki/COVID-19_pandemic_lockdowns> (2020).

8 IHME. COVID-19 Projections, <https://covid19.healthdata.org/> (2020).

9 Cleveland, W. S. Robust locally weighted regression and smoothing scatterplots. J Am Stat Assoc 74, 829–836 (1979).

10 Cox, N. J. Speaking Stata: Smoothing in various directions. Stata Journal 5, 574–593 (2005).

11 Delen, D., Eryarsoy, E. & Davazdahemami, B. No Place Like Home: Cross-National Data Analysis of the Efficacy of Social Distancing During the COVID-19 Pandemic. JMIR public health and surveillance 6, e19862, doi:10.2196/19862 (2020).

12 Wu, J. T., Leung, K. & Leung, G. M. Nowcasting and forecasting the potential domestic and international spread of the 2019-nCoV outbreak originating in Wuhan, China: a modelling study. Lancet, doi:10.1016/S0140-6736(20)30260-9 (2020).

13 Li, R. et al.. Substantial undocumented infection facilitates the rapid dissemination of novel coronavirus (SARS-CoV-2). Science 368, 489–493, doi:10.1126/science.abb3221 (2020).

14 Lasry, A. et al.. Timing of Community Mitigation and Changes in Reported COVID-19 and Community Mobility - Four U.S. Metropolitan Areas, February 26-April 1, 2020. MMWR Morb Mortal Wkly Rep 69, 451–457, doi:10.15585/mmwr.mm6915e2 (2020).

15 Gatto, M. et al.. Spread and dynamics of the COVID-19 epidemic in Italy: Effects of emergency containment measures. Proceedings of the National Academy of Sciences of the United States of America, doi:10.1073/pnas.2004978117 (2020).

16 Union, E. Single market: A single internal market without borders, <https://web.archive.org/web/20190527230316/https://europa.eu/european-union/topics/single-market_en > (2019).

